# A Compartmental Epidemic Model Incorporating Probable Cases to Model COVID-19 Outbreak in Regions with Limited Testing Capacity

**DOI:** 10.1101/2020.07.30.20165282

**Authors:** A. Hasan, Y. Nasution

## Abstract

We propose a new compartmental epidemic model taking into account people who has symptoms with no confirmatory laboratory testing (probable cases). We prove well-posedness of the model and provide an explicit expression for the basic reproduction number (ℛ_0_). We use the model together with an extended Kalman filter (EKF) to estimate the time-varying effective reproduction number (ℛ_*t*_) of COVID-19 in West Java province, Indonesia, where laboratory testing capacity is limited. Based on our estimation, the value of ℛ_*t*_ is higher when the probable cases are taken into account. This correction can be used by decision and policy makers when considering re-opening policy and evaluation of measures.

## 1. Introduction

Since declared as pandemic by WHO in early of March 2020, the coronavirus disease (COVID-19) has been spread to 216 countries by July 2020. At the end of July 2020, the virus has infected more than 17 milion people (confirmed) and caused more than 650 thousand deaths [1]. Particularly in Indonesia, the number of confirmed positive infection is over 100 thousand cases with over 5,000 deaths [2]. In order to prevent and control the outbreak, in early of April 2020, the Indonesian government began to conduct a Large-Scale Social Restriction (LSSR) at several regions with severe outbreak cases, such as at Jakarta and West Java Province [3]. LSSR includes measures such as closing public places, restricting public transport, and limiting travel to and from restricted regions. This regulation is being controlled by each of local government. To assess the duration of LSSR, local government relies on the estimation value of the time-varying effective reproduction number (ℛ_*t*_) at each region [4]. The government officials use sequential Bayesian method based on the discrete SIR model presented in [5] and [6]. The method requires daily confirmed infection data as inputs to estimate ℛ_*t*_.

Based on CDC case definitions, however, there are two classifications of COVID-19 case, namely probable cases and confirmed cases [7]. A person can be classified into probable case if he or she

- *meets clinical criteria AND epidemiologic evidence with no confirmatory laboratory testing performed for COVID-19*,
- *meets presumptive laboratory evidence AND either clinical criteria OR epidemiologic evidence*,
- *meets vital records criteria with no confirmatory laboratory testing performed for COVID-19*.

Details regarding clinical criteria, epidemiologic evidence, and vital records criteria can be found in [7]. Probable case is quite similar to Patient under Surveillance (*Pasien Dalam Pengawasan or PDP*) based on the Indonesian ministry of health [8]. In many provinces in Indonesia, the number of probable cases far exceed the number of confirmed cases, especially in the beginning of the pandemic. For example, Figure 1 shows a much larger number of probable cases in West Java province in Indonesia compared to active/confirmed cases. Limited laboratory testing and the existence of lag time between testing and result can be the cause of this situation [9]. Our aim is to estimate the time-varying effective reproduction number ℛ_*t*_ by utilizing daily probable case data together with daily confirmed case data. To this end, we propose a Susceptible-Probable-Infectious-Recovered (SPIR) model with four compartments as shown in Figure 2.

**Figure 1:**
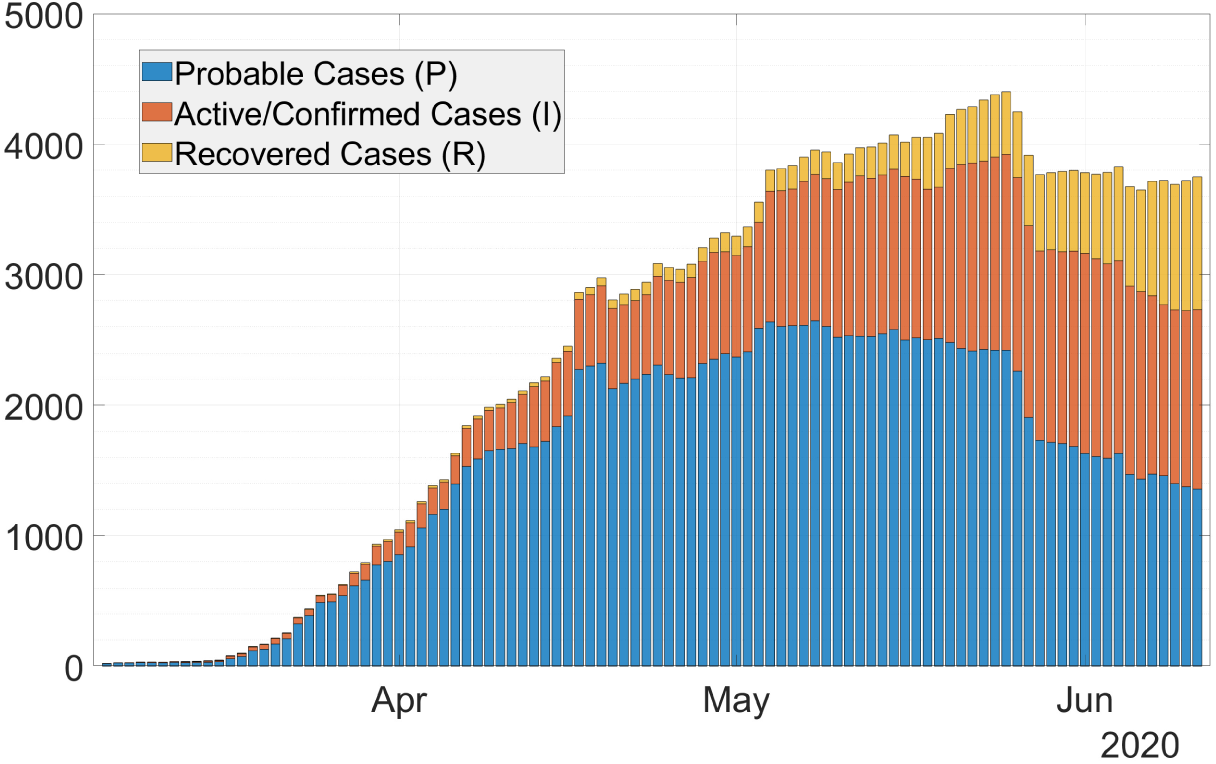
Stacked-bar of COVID-19 cases in West Java, Indonesia.

**Figure 2:**
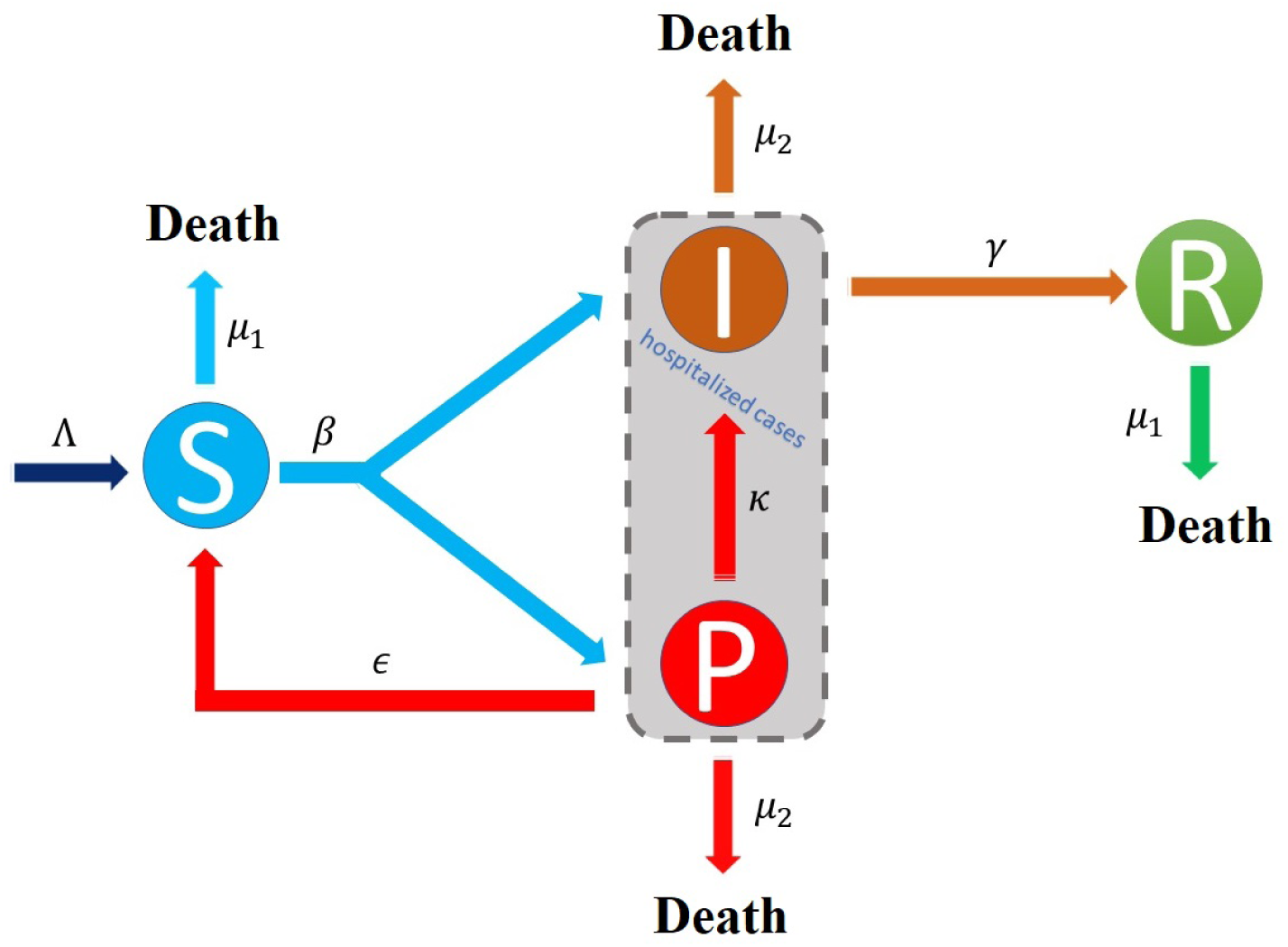
SPIR model is proposed to model probable cases of COVID-19 transmission process.

## 2. Model description

### 2.1 Susceptible-Probable-Infectious-Recovered (SPIR) model

The SPIR model consists of four ordinary differential equations (ODEs), describing the evolution of the population in each stage over time, and is given by:

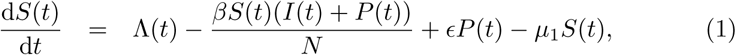

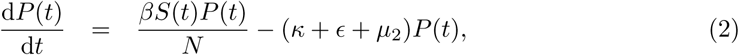

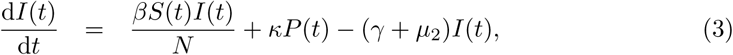

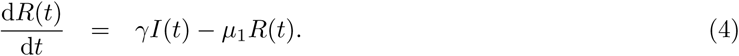

Here, *S, P, I, R* denote susceptible case, probable case, active/confirmed case, and recovered case, respectively. The system satisfies

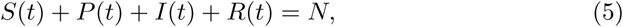

expressing in mathematical terms the constancy of population *N*. We assume the natality Λ(*t*) compensate for the death, i.e.,

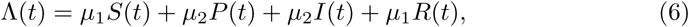

where *µ*_1_ and *µ*_2_ denote the death rates. In our model, we assume *µ*_2_ *> µ*_1_ since infected individuals die at faster rate. If *µ*_1_ = *µ*_2_ = *µ*, then the natality is constant, i.e., Λ = *µN*. The remaining parameters include the infection rate *β*, the negative testing rate *ϵ*, the positive testing rate *κ*, and the recovery rate *γ*. Among these parameters, *β* could be considered as the most important parameter, which quantifies the transmission of the virus. *β* could also be defined as the average number of contacts per person per time multiplied by the probability of disease transmission. Thus, in practice this parameter is time-varying due to intervention. Remark that in order to simplify the analysis, the infection rate *β* in our model goes equally to *I* and *P*. However, if a Polymerase Chain Reaction (PCR) testing shows negative result, a person from compartment *P* can go back to compartment *S*. This is modelled by the negative testing rate *ϵ*.

### 2.2 Well-posedness of the SPIR model

We establish Theorem 1 and Theorem 2 to ensure the mathematical and biological well-posedness of the SPIR model (1)-(4).

**Theorem 1**. *For S*(0), *P* (0), *I*(0), *R*(0) *∈* ℝ, *there exists t*_0_ *>* 0 *and continuously differentiable functions S*(*t*), *P* (*t*), *I*(*t*), *R*(*t*) : [0, *t*_0_) *→* ℝ, *such that* (*S*(*t*), *P* (*t*), *I*(*t*), *R*(*t*)) *satisfies* (1)*-*(4).

*Proof*. The Jacobian of (1)-(4) is linear with respect to *S*(*t*), *P* (*t*), *I*(*t*), and *R*(*t*). Thus, the system is locally Lipschitz. From Picard-Lindelöf theorem [10], there exists a unique solution for the system (1)-(4), hence completes the proof.

**Theorem 2**. *Let us assume that S*(0), *P* (0), *I*(0), *R*(0) *≥* 0. *If the unique solution of* (1)*-*(4) *exists on the interval* [0, *t*_0_] *for t*_0_ *>* 0, *then the functions* (*S*(*t*), *P* (*t*), *I*(*t*), *R*(*t*)) *will remain positive and bounded ∀t ∈* [0, *t*_0_], *i*.*e*., *the system* (1)*-*(4) *is a dynamical system on the compact set*

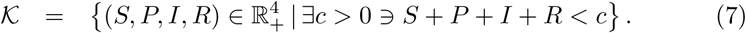

*Proof*. Solution of (2) is given by

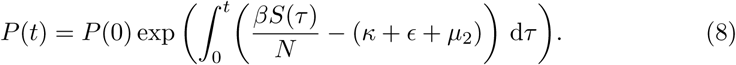

Thus, for *P* (0) *≥* 0 we have *P* (*t*) *≥* 0. Furthermore, solution for (1) is given by

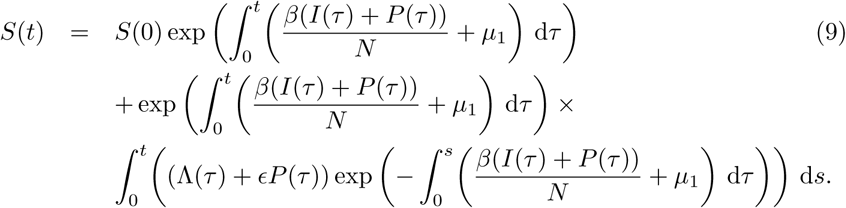

□

Since Λ(*t*) *≥* 0 and *P* (*t*) *≥* 0, then *S*(*t*) *≥* 0. Using similar approach, we can prove *I*(*t*), *R*(*t*) *≥* 0. Boundedness follows from the fact that *S*(*t*)+*P* (*t*)+*I*(*t*)+ *R*(*t*) = *N*.

### 2.3 Basic reproduction number

The SPIR model has a Disease-Free Equilibrium (DFE) state, which is given by (*S*^*\**^, *P*^*\**^, *I*^*\**^, *R*^*\**^) = (*N*, 0, 0, 0). Following Lemma 1 in [11], we define:

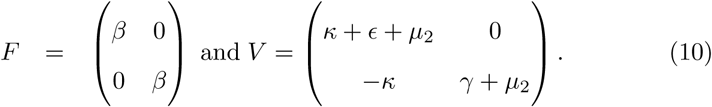

According to [12], the next generation matrix is defined as *FV* ^*−*1^ and the basic reproduction number is defined as the spectral radius of the next generation matrix, i.e.,

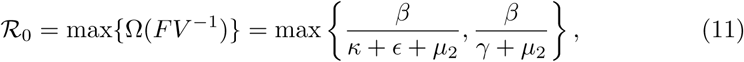

where Ω denotes the eigenvalue of the next generation matrix *FV* ^*−*1^. Remark that, in practice *β* = *β*(*t*) due to intervention. Thus, considering the number of susceptible individual decline over time [13], the time-varying effective reproduction number is given by:

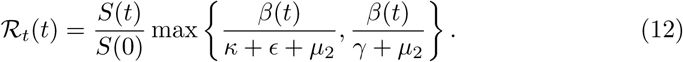

In the next section, we use an extended Kalman (EKF) filter to estimate the infection rate *β*(*t*), and thus estimate the time-varying effective reproduction number ℛ_*t*_(*t*).

## 3. Estimation of the time-varying effective reproduction number ℛ_*t*_

Since Λ(*t*) = *µ*_1_*S*(*t*) + *µ*_2_*P* (*t*) + *µ*_2_*I*(*t*) + *µ*_1_*R*(*t*), the SPIR model (1)-(4) can be written as follow

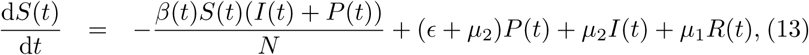

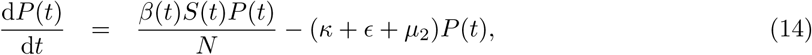

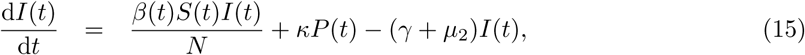

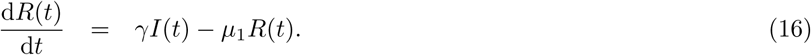

### 3.1 Discrete-time stochastic augmented SPIR model

Discretizing the SPIR model (13)-(16) using forward Euler method and augmenting the infection rate as a sixth state variable, we obtain the following discrete-time stochastic augmented SPIR model:

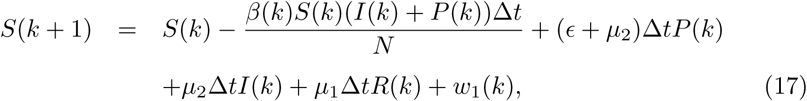

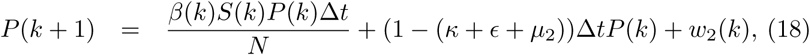

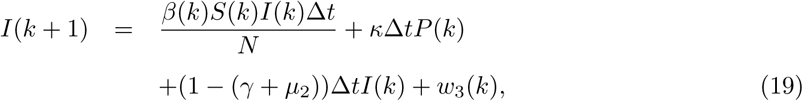

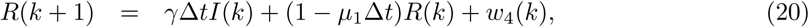

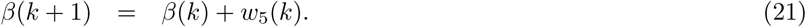

Here, we add noise ***w***(*k*) = (*w*_1_(*k*) *w*_2_(*k*) *w*_3_(*k*) *w*_4_(*k*) *w*_5_(*k*))^τ^ as uncertainty in the model and is assumed to be white noise Gaussian and uncorrelated. Furthermore, we assume the infection rate *β*(*k*) is a piece-wise continuous function with jump every one day. Thus, in one day the value of *β*(*k*) is constant and (21) is satisfied. Augmenting a parameter as a new state variable is a common technique when estimating a parameter using EKF (see page 422 in [14]).

### 3.2 Extended Kalman filter (EKF)

In this section, we use EKF to estimate the state variables in (17)-(21) dynamically. To simplify the presentation, we define an augmented state vector

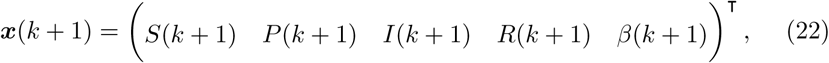

and as such, the discrete-time stochastic augmented SPIR model (17)-(21) can be written as follows

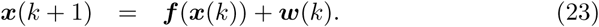

Let us denote 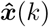 as an estimated vector state from the EKF. Applying first-order Taylor series expansion to ***f*** at 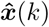, we obtain 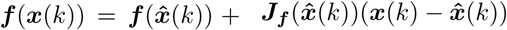, where 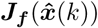 is the Jacobian matrix of ***f***, given by

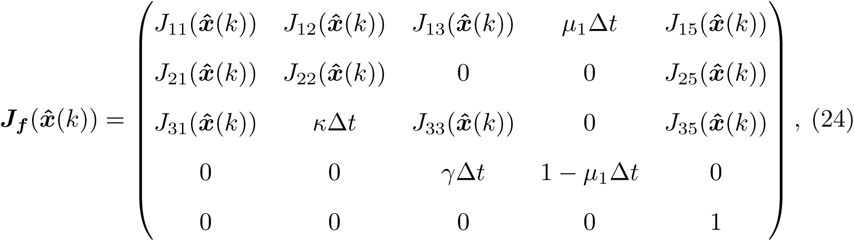

where

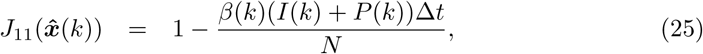

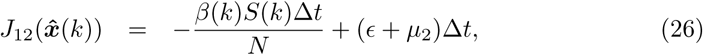

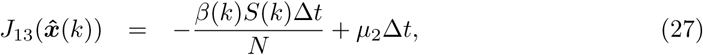

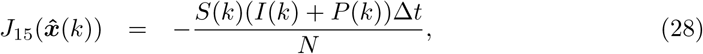

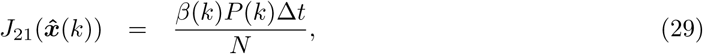

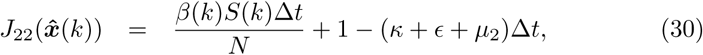

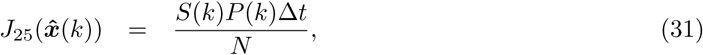

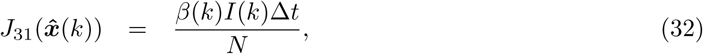

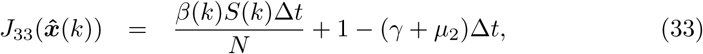

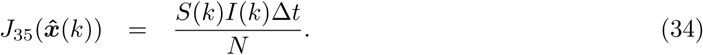

The EKF has two main tuning parameters: the process covariance matrix ***Q***_*F*_ and the observation covariance matrix ***R***_*F*_. Detail procedures of implementing this method can be found in [15]. Note that, the main purpose of EKF is used as real-time data fitting. Thus, the tuning parameters are chosen such that the Relative Root Mean Square Error (RRMSE) between the data and the estimate is sufficiently small. The RRMSE for each variable *X*_*i*_ is defined as

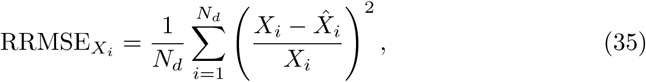

where *N*_*d*_ is the number of observed days. Here, *X*_*i*_ *∈ {S*(*i*), *P* (*i*), *I*(*i*), *R*(*i*)*}* and 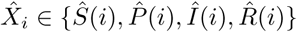 denote the data and the estimate, respectively.

## 4. Case study: West Java province in Indonesia

We use the discrete-time stochastic augmented SPIR model together with the EKF to estimate the time-varying effective reproduction number *R*_*t*_ in West Java province (population 48 million) in Indonesia. All data sets and MATLAB code are available on GitHub (https://github.com/agusisma/covidPDP). To simplify the model, the parameters are assumed to be constant and are obtained from clinical information, such as case-fatality-rate (CFR), infectious time *T*_*i*_, and life expectancy *T*_*l*_, and are given by:

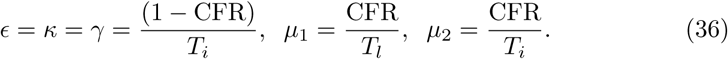

The negative testing rate *ϵ*, the positive testing rate *κ*, and the recovery rate *γ* are assumed to be equal after confirmed through PCR testing. The data are provided in Table 1. The tuning parameters for the Kalman filter are chosen as ***Q***_*F*_ = diag(10 10 10 5 0.2) and ***R***_*F*_ = diag(100 10 5 1), respectively. Figure 3 shows results from dynamic data fitting using EKF. As can be seen from this figure, the discrete-time stochastic augmented SPIR model together with the EKF are able to model the transmissions of COVID-19 accurately.

**Table 1:**
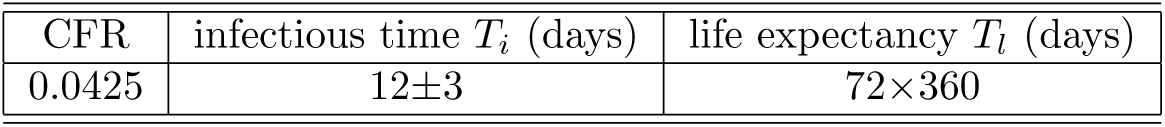
Data used to calculate model parameters.

**Figure 3:**
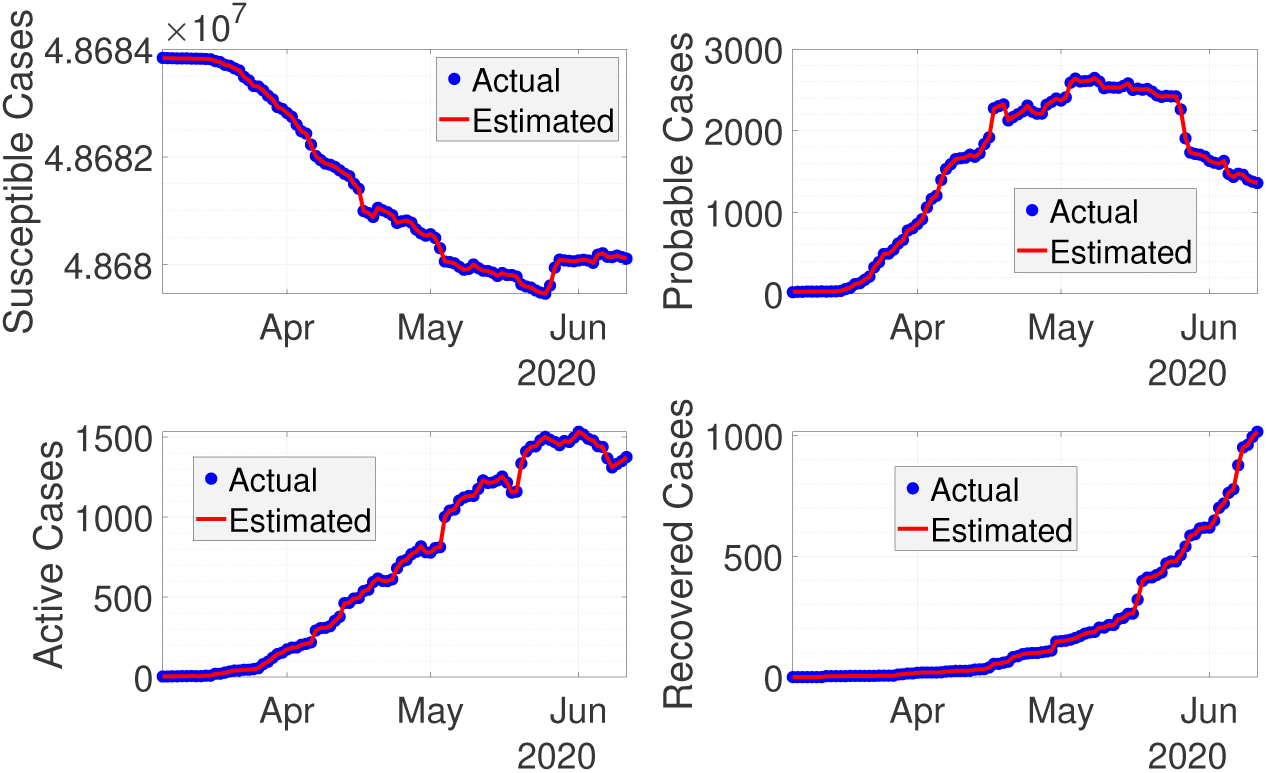
Real-time fitting using extended Kalman filter.

The 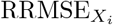 using the chosen tuning parameters ***Q***_*F*_ and ***R***_*F*_ are given in Table 2, where the error is reasonably small.

**Table 2:** RRMSE between reported data and estimated data using EKF.

| RRMSE$_{X_i}$ | | | | |
| --- | --- | --- | --- | --- |
| S | P | I | R | Total |
| 4.1789e-16 | 6.6468e-04 | 1.3062e-06 | 2.2014e-06 | 6.6819e-04 |

The estimated ℛ_*t*_ can be seen from Figure 4. Here, the value of ℛ_*t*_ is higher when the probable cases are taken into account. The difference between the estimated ℛ_*t*_ with and without probable cases becomes smaller once the rate of the probable cases decreases.

**Figure 4:**
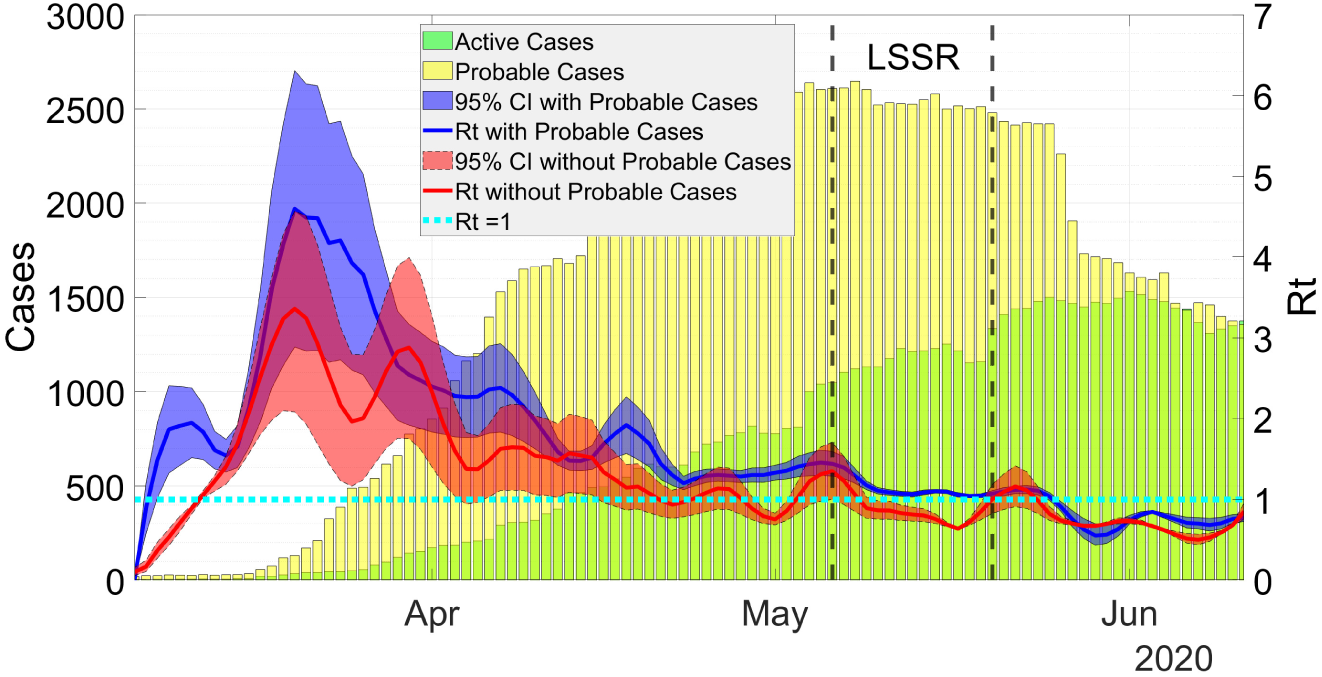
Estimation of ℛ_*t*_ with and without probable cases.

On May 6, the government of West Java province enact a Large-Scale Social Restriction (LSSR). The decision was taken based on the evaluation of ℛ_*t*_, which according to WHO need to be done if ℛ_*t*_ *>* 1. The restriction can be relaxed if ℛ_*t*_ *<* 1 for fourteen consecutive days. Since the government officials estimated ℛ_*t*_ without considering probable cases, the restriction was relaxed on May 20 when ℛ_*t*_ *<* 1 for two weeks. Taking probable cases into account, our estimation shows that during the period of LSSR, the estimated value of ℛ_*t*_ is still above 1. Thus, the LSSR need to be extended rather than relaxed. The effect of LSSR, where ℛ_*t*_ < 1, can be seen after two weeks (May 24) since there are delays in infection confirmation.

## 5. Conclusion

A new compartmental epidemic model taking probable cases into account has been presented in this paper. The model, called the SPIR model, consists of four compartments: Susceptible (S), Probable (P), Active (I), and Recovered (R). The model is used to estimate the time-varying effective reproduction number ℛ_*t*_ in West Java province in Indonesia. To this end, we apply an EKF to a discrete-time stochastic augmented SPIR model. Numerical simulations show that when probable cases are taking into account, the value of ℛ_*t*_ is significantly higher. This results can be used to inform policy makers when deciding to loosen or tighten the measures. In general, our model and approach can be used in regions with limited testing capacity.

## Data Availability

Link to data and codes are provided in the manuscript.

https://github.com/agusisma/covidPDP

## Acknowledgement

We would like to thank Dr. Rangga Ganzar Noegraha who proofreads the paper and Muhammad Fakhruddin who provides Figure 2.

